# Boosting Chinese healthcare service providers’ utilization of behavioural and pharmacotherapy interventions for cigarette smoking cessation by ‘WeChat WeQuit’ program

**DOI:** 10.1101/19012682

**Authors:** Yanhui Liao, Yunfei Wang, Zhenzhen Wu, Yuhang Liu, Chudong Wang, Jinsong Tang

## Abstract

**Introduction:** In China, standard smoking cessation practices are rarely used by healthcare service providers (HSPs). WeChat, a popular social media app, has been widely used in China.

**Methods:** In this single-blind, randomized trial, undertaken in China with 8-week interventions and follow-up to 52 weeks, about 2,200 providers from different health care settings will be randomly selected to the intervention or control group. This trial will be conducted in China between June 2018 to October 2019. The intervention group will receive regular smoking cessation training program messages by the professional team to 8 weeks and follow to 52 weeks. A hard copy of the manual will be sent to each provider from the intervention group by mail after randomization. The Control group will only communicate by themselves and receive thanks messages for 8 weeks, and follow-up to 52 weeks. The trial will be carried out in two phases. The first phase is the pilot study (n=200, 8-week intervention and follow-up to 16 weeks) and the second is the main study (n=2000, 8-week intervention and follow-up to 52 weeks). The primary outcome measure will be the utilization rate of behavioural and pharmacotherapy interventions for smoking patients from 8 to 52 weeks. This trial is registered at ClinicalTrials.gov (number NCT03556774).

**Conclusions:** This program will be the first evidence-based educational program in smoking cessation designed specifically for the improvement of Chinese HSPs’ utilization of behavioural and pharmacotherapy interventions for cigarette smoking cessation in health care settings by the ‘WeChat WeQuit’ program.

**Implications:** This protocol may show that ‘WeChat WeQuit’ training program will be effective in increasing the provision of effective tobacco cessation interventions by Chinese-speaking HSPs, especially therapists, to patients with cigarette smoking, which will provide valuable insights into bridging the gap between need and services for smoking cessation in China. Overall, we believe this program will be likely to have very substantial public health benefits if it would provide a widely accessible and efficacious smoking cessation information for Chinese HSPs.

## Introduction

With over 300 million smokers, cigarette smoking remains a major public health concern in China^1^. In 2015, there were 933.1 million daily smokers in the world, and 6.4 million deaths (11.5% of global deaths) were attributable to cigarette smoking worldwide. Over three quarters of deaths attributable to smoking were in men, and 52.2% took place in four countries (China, India, the USA, and Russia) with China having the highest proportion^2^. Smoking cessation remains the single most effective prevention for lung cancer and other smoking related health issues^3^. However, long-term smoking cessation rates are very low (less than 10% quit rate)^4^. Our previous “Happy Quit” program in China (mobile phone-based text messaging interventions) showed only approximately 6% continuous abstinence rate at 24 weeks ^5,6^.

In China, male physician smoking prevalence is high with few former smokers, and standard smoking cessation practices are rarely provided by HSPs^7-9^. Evidence showed that smoking HSPs were less likely to ask about smoking status and advise smoking patients to quit, and physicians with higher perceived quality of their training in smoking cessation methods led to greater utilization of evidence-based cessation interventions^10^. Thus, health-care professionals should take more responsibility for providing smoking cessation services that are readily available, effective and cheap. Taking the National Health Service (NHS) Stop-Smoking Service (SSS), a national evidence-based, effective, and incredibly cost-effective training program, for example, “Specialist” practitioners in the SSS reported higher success quit rates than “community” practitioners^11^. Also clients who set a quit date with the SSS are more likely to maintain long-term abstinence^12^. The SSS training program shows greater improvements in successful quit rates and provides a cost-effective way of increasing the number of people saved by the SSS^13^. However, the availability of smoking cessation training programs in China is extremely limited, and the majority of cessation attempts end in relapse^14^. Insufficient smoking cessation training programs for HSPs, and therefore insufficient smoking cessation services, would be the most important contributing factor to the low cessation rates reported in China^7^. Furthermore, clinical studies on smoking cessation remain extremely inadequate in China ^15^. A sample from 21 cities in China reported that almost half of smokers intended to quit. However, the prevalence of smoking cessation among those urban-based smokers was only about 10% ^16^. There is an urgent need to improve the utilization of behavioural and pharmacotherapy interventions and to reach underserved populations in China.

Smoking cessation interventions delivered by digital media, such as text messages, WeChat, Facebook, e-mails, web pages, and digital TV can be made widely available to those treatment-seeking smokers for little more than the cost of designing and testing the intervention. Previous trials on digital and social media Interventions for smoking cessation have documented the long-term treatment effects^17,18^. For example, the Happy Ending, with 12 months follow-up, demonstrated the efficacy of the fully automated digital multi-media smoking cessation intervention ^18^. The txt2stop, a mobile phone text messaging smoking cessation program, also showed significantly improved smoking cessation rates at 6 months^19^.

Since its first release in 2011, WeChat (Chinese: 微信; pinyin: Wēixìn; literally: “micro-message”) has become the most popular social media app in China (https://en.wikipedia.org/wiki/WeChat). It has been widely used, either by individuals or groups, among HSPs and patients to promote human health. With the rapid increase of WeChat users during the past several years, most of the health care settings have built WeChat public service platforms to provide instant medical service^20^. There is evidence to show that WeChat-based medical services can improve the quality of care and treatment efficacy. Take smoking cessation services, for example. A sample of eighty-eight patients with coronary heart disease who underwent percutaneous coronary intervention (PCI) were randomly selected to WeChat Groups with smoking cessation intervention or without intervention. This showed that using the WeChat Group-based smoking cessation improved these patients’ quit rates^21^. Two similar smoking cessation studies in China showed that combination therapy of varenicline with a WeChat-based service platform is better than varenicline alone for patients with chronic obstructive pulmonary disease (COPD)^22,23^.

In order to minimize the huge gap between the shortage and demand for smoking cessation training programs and smoking cessation services, we propose this ‘WeChat WeQuit’ smoking cessation training program for Chinese-speaking HSPs to increase the utilization of behavioural and pharmacotherapy interventions for cigarette smoking cessation.

## Methods

### Setting and sample

This large, simple randomized trial of boosting Chinese HSPs’ utilization of behavioural and pharmacotherapy interventions for cigarette smoking cessation by ‘WeChat WeQuit’ program will be conducted using WeChat-based messaging in any region or city of China. Eligible Chinese-speaking HSPs (anticipated participants: 2200, with 200 participants in the pilot study and 2000 participants in the main study) will be randomly selected to the intervention or control groups in a 1:1 ratio.

Eligibility: Chinese-speaking HSPs who are living in China.

Inclusion Criteria

1. Chinese-speaking HSPs
2. Know how to use WeChat
3. Use WeChat on a daily basis
4. Willing to provide informed consent to participate in the study

Exclusion Criteria:

1. Non-Chinese speakers
2. Not HSPs
3. Do not use WeChat
4. Unwilling to participate in the study

### Patient Involvement

Patients were not directly involved in this study.

### Research question/objective

#### Overall objective

this research is designed to assess whether ‘WeChat WeQuit’ smoking cessation training program will increase Chinese HSPs’ utilization of behavioural and pharmacotherapy interventions for cigarette smoking cessation or not.

### Objectives of the pilot study

1. Overall design considerations in developing ‘WeChat WeQuit’ program are aimed at increasing HSPs’ utilization of behavioural and pharmacotherapy interventions for cigarette smoking cessation in China.
2. Examine the content of messages that will be sent by WeChat in this study:
  A. intervention-group messages
  B. control-group messages
3. Quit message types and number of messages per quitting stage.
4. How often messages will be received and read by HSPs.
5. HSPs’ views on the intervention messages and their delivery.
6. The proportion of HSPs and smokers who are contactable at follow-up and who provide outcome variables.
7. Sample size assessment for the main study.
8. The knowledge about behavioural and pharmacotherapy interventions for smoking cessation before and after 8 weeks training.
9. Preliminarily examination of the feasibility and effectiveness of ‘WeChat WeQuit’ program in China.

### Objectives of the main study

1. The primary outcome will be the utilization rate (i.e. rate=smokers treated divided by smokers seen) of behavioural and pharmacotherapy interventions by HSPs for smoking patients from 8 to 52 weeks. The number of smokers seen by each HSP, as well as the number of smokers given treatment by each HSP will be measured. The utilization rate will be recorded by each HSP.
2. The secondary outcome measure will be the proportion of smoking patients with continuously self-reported abstinence at week 1, 4, 12 and 24 follow-ups. The association between particular behavioural change techniques or medications by HSPs and higher rates of successful quitting will be measured as well.
3. A third outcome will explore potential wider influence of the ‘WeChat WeQuit’ training program in China.

Previous studies have shown that providing behavioural and pharmacotherapy interventions for cigarette smoking patients was rare among Chinese HSPs. Providing the ‘WeChat WeQuit’ cessation program for Chinese HSPs may increase the utilization of behavioural and pharmacotherapy interventions for cigarette smoking cessation and increase abstinence rates. We will test the hypothesis that the ‘WeChat WeQuit’ training program will improve Chinese HSPs’ utilization of behavioural and pharmacotherapy interventions for cigarette smoking cessation and increase the likelihood of cessation rates in smoking patients.

### Recruitment

We will advertise this service in the hospitals, private clinics, pharmacies and online (e.g., WeChat, websites, such as http://www.dxy.cn/, QQ). Potential participants will register their interest by sending WeChat messages, text messages or making a call. Research assistants then will contact respondents to assess eligibility and collect baseline data.

### Interventions and procedures

In this single-blind, randomized trial, about 2200 HSPs will be randomly selected (by randomizeR, https://CRAN.R-project.org/package=randomizeR) to 8-week intervention (behavioural and pharmacotherapy interventions for cigarette smoking patients to help them quit smoking) groups or control groups that only communicate without any standard smoking cessation practices related messages, and follow-up to 52 weeks. A research assistant will assign participants to either intervention or control group. Participants, investigators and other research personnel will be masked to treatment allocation. This trial will be carried out in two phases. The first phase is the pilot study (see **Figure 1**. Flowchart for the pilot study) and the second is the main study (see **Figure 2**. Flowchart for the main study).

**Figure 1.**
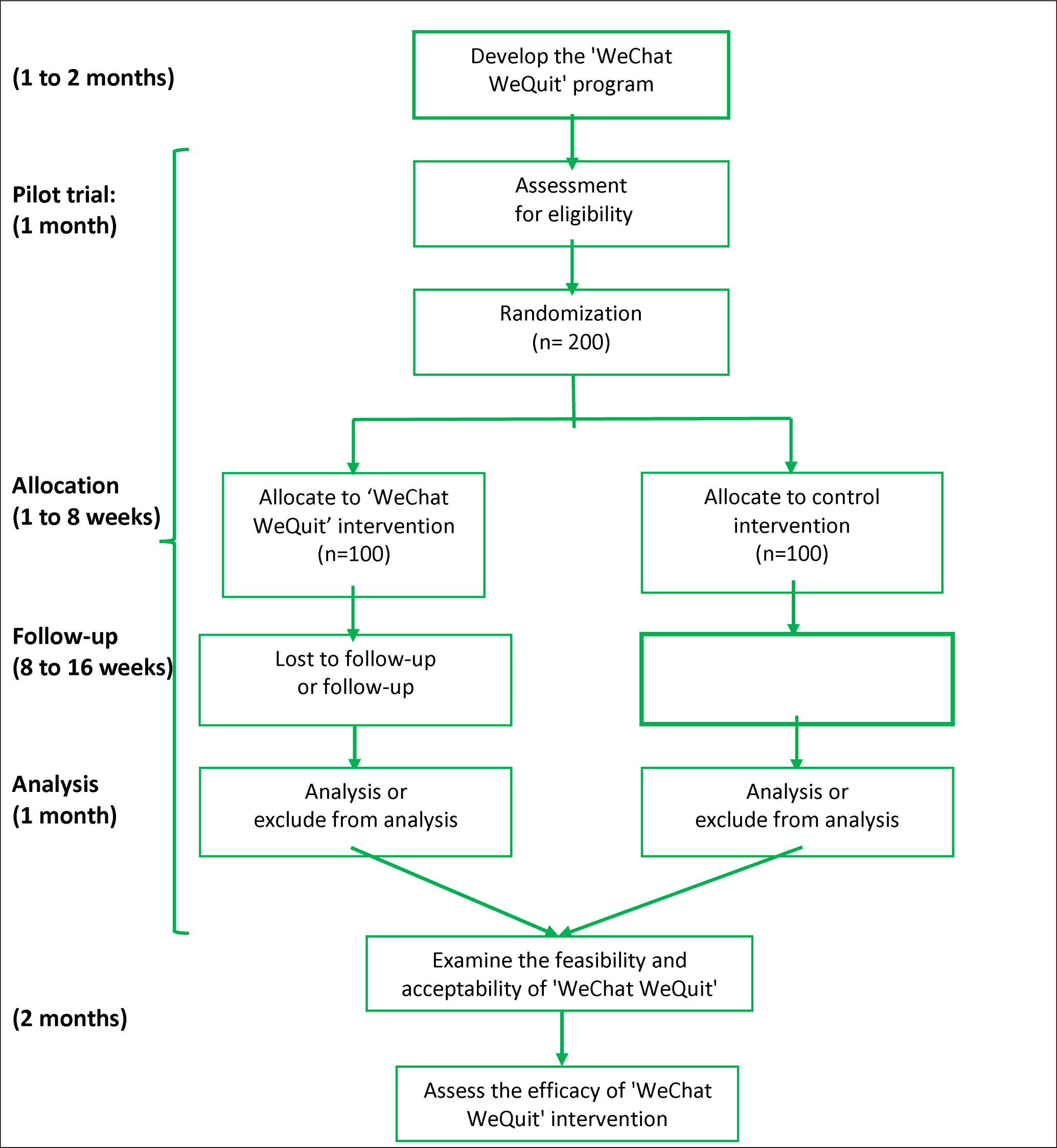
Flowchart for the pilot study.

**Figure 2.**
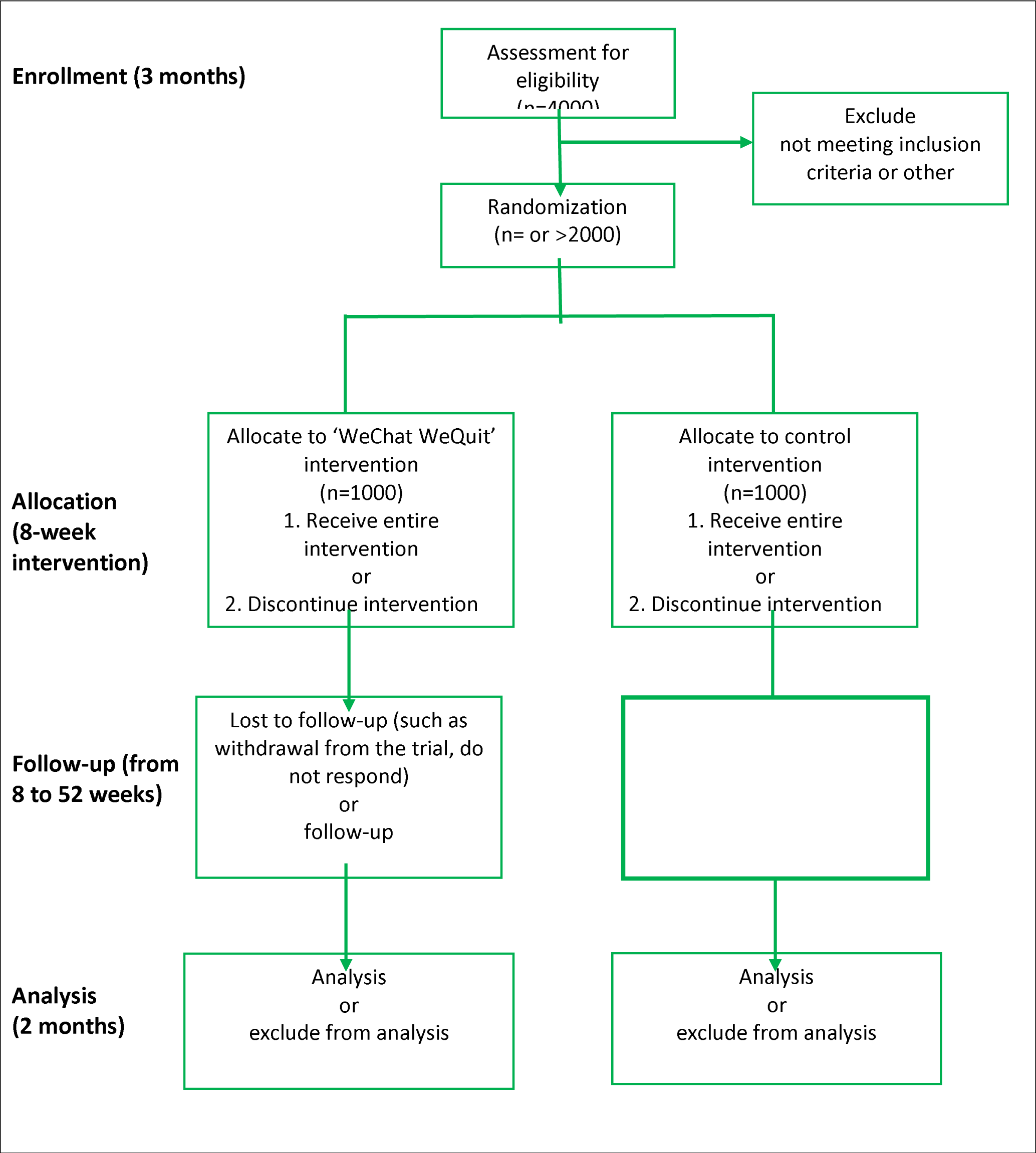
Flowchart for the main study.

### A. The pilot study

The pilot study is to develop a ‘WeChat WeQuit’ program that aims at increasing HSPs’ utilization of behavioural and pharmacotherapy interventions for cigarette smoking cessation in China. The pilot study is to assess the content of both intervention group and control group messages that will be sent by WeChat in this study; quit message types and number of messages per quitting stage; how often messages will be received and read by HSPs; HSPs’ views on the intervention messages and their delivery; the proportion of HSPs and smokers who are contactable at follow-up and who provide outcome variables; sample size assessment for the main study; the knowledge about behavioural and pharmacotherapy interventions for smoking cessation before and after 8 weeks. The basis of the selection of questions on ‘knowledge’ was mainly based on *clinical practice guideline for treating tobacco use* and *dependence*^24^; and preliminary examination of the feasibility and effectiveness of the ‘WeChat WeQuit’ program in China. A total of 10% of the whole sample size (about 200 participants) will be conducted during pilot study.

### Develop the ‘WeChat WeQuit’ program

Development of behavioural and pharmacotherapy interventions messages will be mainly based on clinical practice guidelines for treating Tobacco use and dependence^24^, as well as based on previous studies in other countries, and smokers and smoking cessation professionals in China. The 5 A’S of intervention are shown in **Appendix 1** and the 5 R’S of motivation are shown in **Appendix 2**. It will take approximately one to two months to develop the program.

### Examine the feasibility and acceptability of ‘WeChat WeQuit’

The feasibility and acceptability of the ‘WeChat WeQuit’ program will be examined during the whole period of the pilot study stage. Questions for assessing program acceptability are shown in **Appendix 3**. Examinations will include test-retest reliability and internal consistency of the instruments; acceptability of the message contexts; quit message types and number of messages per quitting stage; how often messages will be received and read by HSPs and HSPs’ views on the intervention messages and their delivery; the proportion of HSPs and smokers who are contactable at follow-up and who provide outcome variables; sample size assessment for the main study; the knowledge about behavioural and pharmacotherapy interventions for smoking cessation before and after 8 weeks; and preliminarily examination of the feasibility and effectiveness of the ‘WeChat WeQuit’ program in China. Knowledge about behavioural and pharmacotherapy interventions for smoking cessation before and after 8 weeks is shown in **Appendix 4**. The utilization rate of interventions for smoking patients is shown in **Appendix 5**.

### Assess the efficacy of ‘WeChat WeQuit’ intervention

The pilot study will be implemented with approximate 200 participants (100 HSPs in the intervention group and 100 HSPs in the control group), and feedback (such as acceptability of messages, context and number of messages/day) from participants will be collected to improve the quality of the ‘WeChat WeQuit’ program. In this single-blind, randomized trial, participants will be selected to either a control group or to a group that receives a smoking cessation educational program. The pilot result of the utilization rate of interventions for smoking patients (see **Appendix 5**) and the assessment of HSPs’ liking and understanding of each week’s Knowledge about behavioural and pharmacotherapy interventions for smoking cessation.

### B. The main study

The flowchart for the main study is shown in **Figure 2**. After examining the feasibility and acceptability of the ‘WeChat WeQuit’ program, this trial will recruit about 2000 HSPs in the main study with a 52-week follow-up. The utilization rate (i.e. rate=smokers treated divided by smokers seen) of behavioural and pharmacotherapy interventions by HSP for smoking patients from 8 to 52 weeks will be assessed and recorded by each HSP. The number of smokers seen by each HSP, as well as the number of smokers given treatment by each HSP will be measured. The proportion of smoking patients with continuously self-reported abstinence at week 1, 4, 12 and 24 follow-ups. This trial will also examine the association between particular behavioural change techniques or medications by HSPs and higher rates of successful quitting. In addition, the main study will explore the potential wider influence of the ‘WeChat WeQuit’ training program in China.

All HSPs will receive WeChat-based questions at the first 8 weeks and at the follow-up 9, 12, 20 weeks until 52 weeks about their smoking cessation services (details see **Appendix 5**) and patients’ quit rates (continuously self-reported smoking abstinence) or smoking status (e.g., how many cigarettes and how many cigarettes reduced per day during the last 4 weeks and past 1 week). They will also receive phone call at 4, 8, 12, 20, 28, 36, 44, and 52-week points.

### Intervention group

Participants who allocate to the intervention group will receive regular smoking cessation training program messages by a professional team. One to six messages will be sent per week for 8 weeks. A hard copy of the behavioural and pharmacotherapy interventions manual will be sent to each HSP by mail after randomization. One to six messages will be sent per month until the end of the 1-year follow-up.

### Control group

Control group participants will not receive any smoking cessation messages by the professional team. They will receive messages thanking them for being in the study and reminding them of the time until the 52-week follow-up. One to six messages will be sent per week for 8 weeks.

### For both groups

Knowledge about behavioural and pharmacotherapy interventions for smoking cessation before and after 8 weeks, the utilization rate (rate = smokers treated divided by smokers seen) of interventions for smoking patients from 8 to 16 weeks (for the pilot study) and from 8 to 52 weeks (for the main study) will be measured for both groups. the proportion of smoking patients with continuously self-reported abstinence at week 1, 4, 12 and 24 follow-ups. Both groups will also be encouraged to record 7-day point prevalence of abstinence and continuous smoking abstinence and to communicate the experience of using behavioural and pharmacotherapy interventions with other HSPs.

### Independent variables

Basic information at base line: We will collect data on gender, age, education levels, marital status, clinical sites, years of practice, types of practice, smoking-cessation training experience, and cigarette smoking characteristics for smoking providers only.

### Outcome measures

1. **Program acceptability:** program acceptability will be measured by questions for assessing program acceptability in **Appendix 3**.
2. **HSPs’ Knowledge:** Knowledge about behavioural and pharmacotherapy interventions for smoking cessation before and after 8 weeks will be on a 10-point scale in **Appendix 4**.
3. **Utilization rate:** the utilization rate of interventions for smoking patients from 8 to 52 weeks will be measured by items from **Appendix 5**.
4. **Smoking-cessation outcomes will be recorded by HSPs**
  - **Nicotine dependence:** nicotine dependence will be assessed by Chinese version of Fagerstrom Test for Nicotine Dependence^25^.
  - **Self-reported continuous smoking abstinence:** no more than five cigarettes smoked in the past week at 4 weeks follow-up and no more than five cigarettes smoked since the start of the abstinence period at 6 months of follow-up.
  - **Point prevalence of abstinence:** not even a puff of smoke, for the last 7 days, at 4, 8, 12, 16, 20 and 24 weeks.
  - **Cigarettes smoked per day:** number of cigarettes smoked per day within the past 4 weeks and the approximate total number of smoked cigarettes within the past 4 weeks

### Sample size and power calculation

The sample size assessment and power calculations are mainly based on the primary outcome of “utilization rate of behavioural and pharmacotherapy interventions for cigarette smoking patients from week 8 to week 52”, as well as the secondary outcome measure of “quit rates”. On the basis of the results of the English National Health Service’s (NHS) Stop-Smoking Services research paper^11^ with a total of 573 specialist practitioners and 466 community practitioners, we estimated that, for assessing the utilization of counselling, 124 HSPs (62 HSPs in each group) are required to have a 80% chance of detecting, as significant at the 5% level, an increase in the primary outcome measure from 58.1% in the control group to 80.7% in the intervention group, for assessing the utilization of advising on stop-smoking medications, 348 HSPs are required to have a 80% chance of detecting as significant, at the 5% level, an increase in the primary outcome measure from 85.9% in the control group to 94.7% in the intervention group. For assessing 4-week biologically verified quit rates, 434 HSPs are required to have 80% chance of detecting as significant, at the 5% level, an increase in the primary outcome measure from 50.4% in the control group to 63.6% in the intervention group. However, a larger sample size will be required to assess 24-week continuous quit rates between two groups.

We then assessed the sample size based on previous randomized controlled trials (RCTs) of smoking cessation programs and Internet interventions, which suggest that quit rates for smokers may be as high as 10% to 20% in the treatment group and as low as 5% in the control group ^26-28^. A conservative estimate of 10 % (treatment) and 5 % (control) self-reported prevalence abstinence at 12- and 24-week follow-up was made. A sample size of 864 will provide a power level of 0.80, and a Type-I error rate of 0.05 to detect these cessation rates in the most conservative outcome of the study self-reported prevalence abstinence. However, considering relatively large drop-offs ^29^, a total sample size of 2200 participants in this study (1100 in each arm) will have more than a 95% chance of detecting a significant difference.

### Data analysis

Statistical analyses will be conducted with R software (https://www.r-project.org/) and SPSS version 22 (IBM Corp. Released 2013. IBM SPSS Statistics for Windows, Version 22.0. Armonk, NY: IBM Corp.). Data will be double checked, and groups with be blinded to statistician. Basic information (including demographic characteristics, cigarette smoking characteristics) at baseline and the overall quality and efficacy of the “WeChat WeQuit” training program (including program acceptability, HSPs’ knowledge and competences, utilization rate,) between study groups using the χ2 (chi square) or Fisher exact test for categorical data and the t-test or Wilcoxon rank-sum test for continuous data will be collected. Smoking-cessation outcomes will be recorded by HSPs. The utilization rate of interventions will be compared between HSPs in the intervention group and the control group using a mixed-effects statistical model. As for smoking status and quit rates, all smoking patients who visited HSPs from this trial will be recorded and analyzed with an “intention-to-treat” analysis. Self-reported continuous abstinence from 1 week to 24 weeks after quit date will be compared between two groups using the χ2. The independent variables will be intervention versus the control condition, assessment points, and covariates identified in preliminary analysis. This model will be estimated using maximum likelihood estimation. All tests will be 2-tailed. A two-sided P<.05 will be used to determine statistical significance.

## Discussion

Since its first release in 2011, WeChat, as a free instant messaging application for smartphones that enables the exchange of text, voice, pictures, videos, and location information between individuals and groups via mobile phone indexes, has become the most popular social media app in China. In addition to its service in communication, library, financial and other fields, it has been widely used to promote human health. Thus, by this ‘WeChat WeQuit’ educational program, Chinese HSPs could easily learn how to utilize behavioural and pharmacotherapy interventions for cigarette smoking patients to stop smoking and prevent relapse. This WeChat-based educational program of smoking cessation may dramatically minimize the huge gap between the shortage and demand for smoking cessation training program and smoking cessation service. According to the National Health Service Stop-Smoking Service (SSS), HSPs with smoking cessation services training would more likely to help smoking patients quit smoking^11^. As the SSS training program shows its efficacy in quitting smoking and provides a cost-effective way of increasing the number of people saved by the SSS^13^, this ‘WeChat WeQuit’ program is very likely to provide a cost-effective way to reach large numbers and achieve a substantial impact.

However, although almost every Chinese HSP has a smartphone and uses WeChat, some participants may not be serious enough in reading messages as a few WeChat users complain about bombardment by excessive information. WeChat users today are constantly bombarded by messages and can easily ignore them. In order to increase the utilization of the program, a hard copy of behavioural and pharmacotherapy interventions manual will be sent to each HSP by mail after randomization.

## Data Availability

No data yet.

## Ethical issues

The protocol was approved by the university ethics committee (The Second Xiangya Hospital of Central South University Review Board, 2018, No. S071) and the studies will be carried out in accordance with the Declaration of Helsinki. At the time of the initial screening interview, the study participants will be required to read the informed consent (confirmation of electronic version of written informed consent will be performed) and may ask any questions they may have about it. At the same time, a researcher will read the consent statement to every HSP during the recruitment process. Each participant will receive an explanation about the aims, importance, procedures, measurements, potential risks and benefits of the study before recruitment. At the time of the initial screening interview and again during the orientation session preceding the study, participants will be required to know about the consent form and may ask any questions they may have about it. Participation will be completely voluntary. If you want to stop this service, you can drop out at any time. Only the investigators can identify the personal information of participants and have access to the final trial dataset. In this single-blind, randomized trial, you will have a 50 % chance to be selected to either a control group or to a group that receives the ‘WeChat WeQuit’ program. If you are in the control group, you are welcome to join in the ‘WeChat WeQuit’ program after the study. Participants will be given contact details of the study coordinator for their future questions and concerns. Thus, all participants will be able to contact the study team for extra advice or suggestions.

## Authors’ contributions

YL developed the ‘WeChat WeQuit’ program and the first draft of this manuscript. JT commented on and revised the manuscript. All authors read and approved the final manuscript.

## Acknowledgments

The authors wish to acknowledge the contributions of Dr. Mei Yang for her assistance in formatting the protocol and Joe Wilson for his assistance in correcting grammatical errors.

## Appendix 1

### The 5 A’S of intervention

1. **ASK - 1 minute** Ask patient to describe their smoking status.
  A. I NEVER smoked or smoked LESS THAN 100 cigarettes.
  B. I stopped smoking more than 2 weeks ago but less than 1 year ago.
  C. I stopped smoking more than 1 year ago.
  D. I smoke regularly/not thinking of quitting in the next 30 days. If B or C, reinforce their decision to quit, congratulate and encourage. If D, document smoking status on their chart. Begin steps below.
2. **ADVISE - 1 minute** Provide clear, strong advice to quit with personalized messages about the impact of smoking on health; urge every tobacco user to quit.
3. **ASSESS - 1 minute** Assess the willingness to make a quit attempt within 30 days.
  - Patient is willing to make a quit attempt in the next 14-30 days
  - Patient is not willing to make a quit attempt (review the 5 R’s)
4. **ASSIST - 3 minutes** Recommend the use of approved pharmacotherapy. Refer to currently available cessation services. **AND/OR** Help the patient develop a quit plan. Provide problem-solving methods and skills for cessation. Provide social support as a part of the treatment. Help patient obtain extra treatment/social support for quitting in the smoker’s environment. Recommend the use of approved pharmacotherapy. Provide self-help smoking cessation materials. Provide relapse prevention.
5. **ARRANGE - 1 minute +** Assess smoking status every visit, reinforce/encourage cessation.

## Appendix 2

### The 5 R’S of motivation

#### Relevance - 1 minute

Ask patient about how quitting may be personally relevant. Encourage the patient to indicate why quitting is personally relevant, being as specific as possible. Motivational information has the greatest impact if it is relevant to a patient’s disease status or risk, family or social situation (e.g., having children in the home), health concerns, age, gender, and other important patient characteristics (e.g., prior quitting experience, personal barriers to cessation). Examples of relevance include:

- Longer and better quality of life
- Extra money
- People you live with will be healthier
- Decrease chance of heart attack, stroke or cancer
- If pregnant, improves chance of healthy baby

#### Risks - 1 minute

Ask the patient about their perception of short-term, long-term and environmental risks of continued use. The HSPs may suggest and highlight those that seem most relevant to the patient. The HSPs should emphasize that smoking low-tar/low-nicotine cigarettes or use of other forms of tobacco (e.g., E-cigarettes, smokeless tobacco, cigars, and pipes) will not eliminate these risks. Examples of risks are:

- Acute risks: shortness of breath, exacerbation of asthma, increased risk of respiratory infections, harm to pregnancy, impotence, infertility.
- Long-term risks: heart attacks and strokes, lung and other cancers (e.g., larynx, oral cavity, pharynx, esophagus, pancreas, stomach, kidney, bladder, cervix and acute myelocytic leukemia), chronic obstructive pulmonary diseases (chronic bronchitis and emphysema), osteoporosis, long-term disability and need for extended care.
- Environmental risks: increased risk of lung cancer and heart disease in spouses; increased risk for low birth weight, sudden infant death syndrome (SIDS), asthma, middle ear disease, and respiratory infections in children of smokers.

#### Rewards - 1 minute

Ask the patient about perceived benefits/rewards for quitting tobacco use. The HSPs may suggest and highlight those that seem most relevant to the patient. Examples of rewards follow:

- Health (self & others)
- Food taste
- Sense of smell
- Feel better
- Example to others
- Additional years of life
- Saving money
- Performing better in physical activities
- Improved appearance including reduced wrinkling/aging of skin and whiter teeth

#### Roadblocks - 3 minutes +

The HSPs should ask the patient to identify barriers or impediments to quitting and provide treatment (problem-solving counseling, medication) that could address barriers. Typical barriers might include:

- Withdrawal symptoms
- Fear of failure
- Sleep problems
- Weight gain
- Lack of support
- Depression and anxiety
- Enjoyment of tobacco
- Limited knowledge of effective treatment options

#### Repetition - 1 minute +

Respectfully repeat the 5 R’s each visit, providing motivation and information. Tobacco users who have failed in previous quit attempts should be told that most people make repeated quit attempts before they are successful.

## Appendix 3

### Questions for assessing program acceptability

1. Overall rating of the program
  A. Like very much
  B. Like somewhat
  C. Neutral
  D. Dislike somewhat
  E. Dislike very much
2. Appraisal of program--likelihood of applying program for smoking patients
  A. Very likely
  B. Somewhat likely
  C. Neutral
  D. Unlikely
  E. Not at all likely
3. Appraisal of program--likelihood of recommending program to other HSPs
  A. Very likely
  B. Somewhat likely
  C. Neutral
  D. Unlikely
  E. Not at all likely
4. I would not have been able to help patients quit without the program
  A. Strongly agree
  B. Agree
  C. Neutral
  D. Disagree
  E. Strongly disagree
5. The program made it easier to help patients quit smoking
  A. Strongly agree
  B. Agree
  C. Neutral
  D. Disagree
  E. Strongly disagree
6. The program disrupted my daily schedule
  A. Strongly agree
  B. Agree
  C. Neutral
  D. Disagree
  E. Strongly disagree
7. Appraisal of WeChat-based messages 7.1 Frequency of reading messages Will be reported on a 10-point scale ranging from 1 (never) to 10 (always).

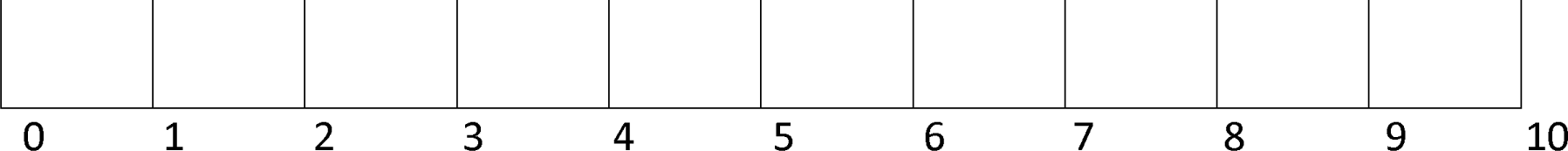 7.2 I received too many messages
  A. Strongly agree
  B. Agree
  C. Neutral
  D. Disagree
  E. Strongly disagree
8. The messages talked about what I was experiencing to learn
  A. Strongly agree
  B. Agree
  C. Neutral
  D. Disagree
  E. Strongly disagree

## Appendix 4

### Knowledge about behavioural and pharmacotherapy interventions for smoking cessation before and after 8 weeks will be on a 10-point scale

**Week 1:** Smoking demographics and the health risks of smoking

**Week 2:** Nicotine dependence, the nature course of smoking, the criteria of nicotine dependence

**Week 3:** Intervention: 5A’s and 5R’s intervention, ABCs (Ask, Brief advice and Cessation support) intervention, SBIRT (Screening, brief intervention, and referral to treatment) intervention

**Week 4:** Quitting preparation: increasing motivation, decreasing triggers, set a quit data, not ready to quit right now, relaxation training

**Week 5:** Psychological and behavioural intervention: Individual counseling and psychotherapy, Group counseling and psychotherapy.

**Week 6:** Pharmacotherapy: nicotine replacement therapy and other medications (e.g. Bupropion and Varenicline)

**Week 7:** Cessation for subpopulation (e.g. women, adolescents, pregnancy and postpartum) and smokers with physical (e.g. cancers, cardiovascular diseases) or mental illness (e.g. schizophrenia, depression, anxiety, drug dependence)

**Week 8:** After quitting: dealing with withdrawal symptoms, addressing weight gain concerns, dealing with lapses, maintaining long-term abstinence

## Appendix 5

### The utilization rate of interventions for smoking patients

1. **The utilization rate of the 5 A’S**
  A. Ask
  B. Advise
  C. Assess: such as
    - Willingness or motivation
    - Preparation
    - Self-efficacy
  D. Assist
    - Set a quit data
    - Recommend cessation program
    - Provide cessation information
    - Discuss or prescribe medications, such as first-line medications:
      – nicotine replacement therapy
      – Bupropion SR
      – Varenicline
  E. Arrange: such as
    - Face to face visits
    - Telephone follow-up
    - Follow-up by WeChat, QQ, email et al.
2. **The overall utilization rate: smokers treated/ smokers seen**
3. **Smoking abstinence rate: the proportion of treated smoking patients with abstinence**

## References

1. Flenaugh EL. Tobacco smoking in China: a pulmonary health crisis. Current opinion in pulmonary medicine. 2019;25(2):188–191.

2. Collaborators GT. Smoking prevalence and attributable disease burden in 195 countries and territories, 1990-2015: a systematic analysis from the Global Burden of Disease Study 2015. Lancet (London, England). 2017;389(10082):1885–1906.

3. Rojewski AM, Zuromski KL, Toll BA. Strategies for smoking cessation among high risk populations to prevent lung cancer. Taylor & Francis; 2017.

4. Messer K, Trinidad DR, Al-Delaimy WK, Pierce JP. Smoking cessation rates in the United States: a comparison of young adult and older smokers. American journal of public health. 2008;98(2):317–322.

5. Liao Y, Wu Q, Kelly BC, et al. Effectiveness of a text-messaging-based smoking cessation intervention (“Happy Quit”) for smoking cessation in China: A randomized controlled trial. PLoS medicine. 2018;15(12):e1002713.

6. Liao Y, Wu Q, Tang J, et al. The efficacy of mobile phone-based text message interventions (‘Happy Quit’) for smoking cessation in China. BMC public health. 2016;16(1):833.

7. Jiang Y, Ong MK, Tong EK, et al. Chinese physicians and their smoking knowledge, attitudes, and practices. American journal of preventive medicine. 2007;33(1):15–22.

8. Smith DR, Zhao I, Wang L. Tobacco smoking among doctors in mainland China: a study from Shandong province and review of the literature. Tobacco induced diseases. 2012;10(1):14.

9. Abdullah AS, Qiming F, Pun V, Stillman FA, Samet JM. A review of tobacco smoking and smoking cessation practices among physicians in China: 1987–2010. Tobacco control. 2013;22(1):9–14.

10. Schoj V, Mejia R, Alderete M, et al. Use of smoking cessation interventions by physicians in Argentina. Journal of smoking cessation. 2016;11(03):188–197.

11. McDermott MS, Beard E, Brose LS, West R, McEwen A. Factors associated with differences in quit rates between “specialist” and “community” stop-smoking practitioners in the english stop-smoking services. nicotine & tobacco research. 2012;15(7):1239–1247.

12. Bauld L, Hiscock R, Dobbie F, et al. English stop-smoking services: one-year outcomes. International journal of environmental research and public health. 2016;13(12):1175.

13. Brose LS, West R, Michie S, McEwen A. Changes in success rates of smoking cessation treatment associated with take up of a national evidencebased training programme. Preventive medicine. 2014;69:1–4.

14. Jiang Y, Elton-Marshall T, Fong GT, Li Q. Quitting smoking in China: findings from the ITC China Survey. Tobacco Control. 2010;19(Suppl 2):i12-i17.

15. Zhang J, Ou JX, Bai CX. Tobacco smoking in China: prevalence, disease burden, challenges and future strategies. Respirology. 2011;16(8):1165–1172.

16. Yang T, Mao A, Feng X, et al. Smoking cessation in an urban population in China. American journal of health behavior. 2014;38(6):933–941.

17. Brendryen H, Drozd F, Kraft P. A digital smoking cessation program delivered through internet and cell phone without nicotine replacement (happy ending): randomized controlled trial. Journal of Medical Internet Research. 2008;10(5).

18. Brendryen H, Kraft P. Happy Ending: a randomized controlled trial of a digital multi-media smoking cessation intervention. Addiction. 2008;103(3):478–484.

19. Free C, Knight R, Robertson S, et al. Smoking cessation support delivered via mobile phone text messaging (txt2stop): a single-blind, randomised trial. The Lancet. 2011;378(9785):49–55.

20. Zhou D, Lu J, Lv X, Zhang L. Research on WeChat medical service model and construction strategy. China Digital Medicine. 2015;10(10):33–35.

21. Li R, Zeng Y. Application of WeChat Group in smoking cessation intervention for middle-aged and young coronary intervention patients who discharged from hospital Chinese Clinical Nursing. 2014(5):389–391.

22. Huang H, Huang X, Lin C. The effect of WeChat guidance combined with varenicline intervention on smoking cessation in patients with chronic obstructive pulmonary disease. Internal Medicine. 2017;12(2):260–262.

23. Yang D, Gu C, Ni L. Evaluation of the efficacy of medication combined with WeChat platform in smoking cessation in patients with chronic obstructive pulmonary disease. Journal of Shanghai Jiaotong University: Medical Science. 2016;36(03):385.

24. Tobacco Tcpgt. A clinical practice guideline for treating tobacco use and dependence: 2008 update: a US public health service report. American journal of preventive medicine. 2008;35(2):158.

25. Huang C-L, Lin H-H, Wang H-H. The psychometric properties of the Chinese version of the Fagerstrom Test for Nicotine Dependence. Addictive Behaviors. 2006;31(12):2324–2327.

26. Swartz L, Noell J, Schroeder S, Ary D. A randomised control study of a fully automated internet based smoking cessation programme. Tobacco control. 2006;15(1):7–12.

27. Muñoz RF, Barrera AZ, Delucchi K, Penilla C, Torres LD, Pérez-Stable EJ. International Spanish/English Internet smoking cessation trial yields 20% abstinence rates at 1 year. Nicotine & tobacco research. 2009;11(9):1025–1034.

28. Cornuz J, Humair J-P, Seematter L, et al. Efficacy of resident training in smoking cessation: a randomized, controlled trial of a program based on application of behavioral theory and practice with standardized patients. Annals of internal medicine. 2002;136(6):429–437.

29. Ramo DE, Thrul J, Delucchi KL, Ling PM, Hall SM, Prochaska JJ. The Tobacco Status Project (TSP): Study protocol for a randomized controlled trial of a Facebook smoking cessation intervention for young adults. BMC public health. 2015;15(1):897.

